# Mechanical feasibility of retrograde trans-aqueduct access to the third ventricle: a human cadaveric and MRI morphometric study

**DOI:** 10.64898/2026.04.14.26348906

**Authors:** Michael Harmer Haines, Stephen Michael Ronayne, Kylie Pickles, David Angus Begg, Peter J. Hurley, Mia Ferraccioli, Patricia Desmond, Nicholas Lachlan Opie

**Affiliations:** Department of Medicine, University of Melbourne, Parkville, VIC, Australia; Department of Anatomy and Physiology, University of Melbourne, Parkville, VIC, Australia; Department of Radiology, University of Melbourne, Parkville, VIC, Australia; Ultra Bionics Pty Ltd., Australia

## Abstract

The third ventricle lies in close proximity to deep brain nuclei, including the subthalamic nucleus (STN) and globus pallidus internus (GPi), that are targeted in proven neuromodulation therapies. This study defines the anatomical pathway via direct and medical imaging analysis; and evaluates the mechanical feasibility of retrograde access to the third ventricle via the cerebral aqueduct using minimally invasive interventional techniques, in cadaveric models.

Evaluation was conducted in three phases using human MRI datasets (n=16; mean age 48.4 years) and cadaveric specimens (n=6; mean age 88.2 years). Phase 1 involved morphometric MRI analysis of the aqueduct and ventricles. Phase 2 tested trans-aqueduct access on cadaver specimens via fluoroscopically guided guidewires and catheters. Phase 3 utilized direct anatomical dissections on cadaver specimens (n=3) to morphometrically measure the third ventricular cavity and its relationship to deep brain nuclei.

Measurements across the sample groups showed a mean aqueduct diameter of 1.6 mm (SD=0.20). Third ventricle dimensions averaged 30.3 mm (anterior-posterior), 18.2 mm (inferior-superior), and 4.8 mm (lateral). Successful access to the third ventricle was achieved in 83% (5/6) of cadaveric specimens via retrograde approach. The system configuration utilised a 0.018″ angled-tip guidewire and 5–6 Fr catheters; the aqueduct accommodated catheter diameters of up to 2.0 mm without strong resistance. The medial margins of the STN and GPi were measured to be approximately 11 and 20 mm, respectively, from the third ventricular centroid.

This research demonstrates that in cadaveric conditions, the trans-aqueduct approach is a mechanically feasible, minimally invasive access pathway to the third ventricle, offering a potential route to the deep brain for therapeutic technologies. Further investigation is required to thoroughly evaluate physiological tolerance, trauma risk, and the long-term implications of intraventricular implantation.

## 1. Introduction

Neuromodulation technologies, including brain computer interfaces (BCI) and deep brain stimulation (DBS) along with emerging techniques such as focused ultrasound (FUS) neuromodulation and transcranial electrical stimulation (tES), offer therapeutic benefits for a range of neurological and psychiatric disorders [1], [2], [3]. Deep cerebral structures are well-established targets for neuromodulation, with target selection guided by the underlying pathophysiology of each condition: the subthalamic nucleus (STN) and globus pallidus internus (GPi) for Parkinson’s disease and dystonia; the ventral intermediate nucleus of the thalamus (Vim) for essential tremor; the centromedian thalamic nucleus for Epilepsy; the anterior limb of the internal capsule or nucleus accumbens for obsessive-compulsive disorder and addiction; and the subcallosal cingulate or ventral capsule for treatment-resistant depression [4], [5], [6], [7], [8], [9]. However, in the case of conventional DBS and BCI, accessing these deep cerebral targets currently necessitates open-brain neurosurgical procedures, posing significant risks to the patient, including trauma, haemorrhage-related risks, infection, and therapeutic benefit through impaired device performance due to sub-optimal electrode positioning or subsequent electrode migration.

Non-invasive neuromodulation methods, such as tES and transcranial FUS, are currently being evaluated for their potential to treat neurological conditions [10], [11], [12], with each showing promising results, however they are limited in their capacity to selectively modulate deep brain targets with high precision due to aberrations and attenuation of both electrical and ultrasonic signals, caused by the skull bone [13], [14]. Consequently, achieving sufficient focal stimulation with these systems often requires higher input power, complex thermal cooling apparatus on the skull, and complicated multi-element arrays, which reduce the practicality of these systems for routine or home-based use.

Emerging minimally invasive approaches that access the brain via the endovascular pathway have shown promise for recording information from large superficial vessels [15], [16], [17], [18], but again, their capabilities remain limited. Deep cortical vessels such as the internal cerebral veins or the basal veins (of Rosenthal), required to deliver devices to within 10mm of the basal ganglia, are small, tortuous, and extremely susceptible to trauma, thrombosis, or occlusion, each of which is likely to be catastrophic to the patient [19]. Additionally, the innate immune response to blood contacting implanted devices promotes a high degree of immunosurveillance, resulting in strong protein adsorption and likelihood of fibrotic encapsulation, potentially impacting its long-term performance [20].

The cerebral ventricular system provides an alternative pathway to deep brain regions which can be accessed via naturally occurring, intrathecal lumens and cavities, without requiring a craniotomy or penetration of delicate neural tissues. For example, the third ventricle, located at the midline between the left and right thalami, is anatomically positioned near several deep brain targets relevant to neuromodulation—namely the STN, GPi, and Vim [21].

Whilst deploying devices in the third ventricle may not provide an advantage for conventional electrical stimulation, development projects are underway to build and evaluate novel devices and systems that can modulate neural populations through intermediate tissue volumes. For instance, by combining systems of ultrasound beam focusing using phased array systems into micro-scale implantable devices, teams such as ours are investigating the ability to reliably modulate neural targets at a separation of distance, from the cerebral ventricles, thereby bypassing the deleterious effects of the skull to deliver precision, steerable, and multifocal neuromodulation. This study aims to evaluate the mechanical feasibility, morphometric analysis, and the gross anatomical arrangement of a **trans-aqueduct approach** to the third ventricle. The trans-aqueduct approach has previously been shown as a viable access route for shunt placement for restoration of cerebrospinal fluid (CSF) flow [22], [23], [24], [25], [26] and researchers have explored the intrathecal route as a pathway to the cortex and the third ventricle via a retrograde third-ventriculostomy approach [27], [28]. By traversing the cerebral aqueduct from the spinal column through the fourth ventricle, access to the third ventricle may be achieved without craniotomy and therefore offer advantages for device implantation and therapy delivery through potential reductions in procedural duration, anaesthetic demand, recovery period, and infection risk. This study investigates the mechanical fundamentals of a retrograde approach to the third ventricle, evaluating the capability of interventional systems to access the cavity, and defining the required anatomical trajectory, for the primary purpose of medical device development. Prior to clinical translation, extensive validation of both the interventional systems and the surgical methodology is required.

## 2. Materials and Methods

### 2.1. Study Endpoints

The primary endpoint of the present study was to describe the mechanical feasibility of retrograde trans-aqueduct catheterisation of the third ventricle in human cadaveric specimens, defined as a catheter tip within the third ventricle, confirmed via contrast ventriculography. Secondary endpoints were also evaluated including: 1. Morphometric analysis of the cerebral aqueduct, third ventricle and related structures via FLAIR MRI datasets; 2. Direct anatomical morphometric measurements on human cadaveric specimens; 3. qualitative evaluation of observed mechanical resistance during catheterisation.

### 2.2. Ethical Approval and Specimen origin

Ten embalmed human head-and-neck specimens were made available through the Body Donor Program, Department of Anatomy and Physiology, The University of Melbourne (Victoria, Australia), (accessed: 06/05/2025). This program requires informed consent from donors and next of kin for the use of donated bodies in research. All specimens were deidentified, with only age, sex, and cause of death supplied to our research team. The anatomical embalming fluid used for preservation contained <2% w/v formaldehyde. A randomly selected subset of six specimens (2 male, 4 female, mean age 88.2 years, SD = 9.1 years) were utilised for trans-aqueduct access evaluation, and a further subset (1 male, 2 female, mean age 92.3 years, SD = 10.9 years) was selected at random from these six specimens to undergo detailed anatomical dissection. Four specimens have been retained for future examinations. This project was approved by the human research ethics committee at the University of Melbourne (Ethics ID: 2024-31208-60686-3).

### 2.3. Anatomical assessment of population MRI data

Sixteen diagnostic fluid attenuated inversion-recovery (FLAIR) 3D MRI datasets were provided by the Dept. of Radiology at the Royal Melbourne Hospital (Victoria, Australia), (accessed: 10/09/2024). Imaging was acquired using a Siemens MAGNETOM Cima.X 3T MRI scanner (Siemens Healthineers, Erlangen, Germany) using a resolution of 0.48 mm per voxel. Patient identity, sex, and health status were fully anonymised prior to data provision. Consequently, specific cerebral pathologies, mild ventricular enlargement, and sub-clinical cerebral atrophy could not be completely excluded from the dataset. Inclusion criteria for the analysis were: age ≥ 18 years; availability of complete 3-dimensional FLAIR MRI imaging encompassing the cerebrum; and image resolution sufficient to delineate the cerebral aqueduct and third ventricular anatomical boundaries. Datasets were to be excluded from analysis if they showed gross anatomical abnormalities (major space-occupying lesions, obvious hydrocephalus, evidence of previous surgical intervention). No datasets were excluded from analysis based on these criteria. The mean age of the patient cohort was 48.4, SD = 17.2 years. The data was evaluated independently by 2 raters using open-source software [29] following a predefined measurement protocol (provided in supplementary data, sections S.2, S.5). This project was exempted of ethics approval by the human research ethics committee at the University of Melbourne (Ethics ID: 2026-36108-79208-1).

#### 2.3.1. Statistical Analysis

Descriptive statistics were reported independently for each rater. Correlation and agreement between the 2 raters were examined using complementary approaches. The linear association between measurements was examined using Pearson’s correlation coefficient and Lin’s concordance correlation coefficient, CCC, was used to quantify agreement with precision and accuracy of measurement. Inter-rater reliability was quantified using the two-way random effects, absolute agreement, Intraclass correlation coefficient, ICC (2,1). Observer agreement was evaluated using the Bland-Altman method which included calculation of the bias, standard deviation of the differences and 95% limits of agreement. Statistical analyses were performed in Visual Studio Code (version 1.109.4) with Python 3 using the Pingouin toolbox (version 0.5.5).

### 2.4. Trans-Aqueduct Access Protocol

Each cadaver specimen was positioned prone (Fig 1, A). This specimen positioning matches the most described patient positioning for retrograde cerebral aqueduct procedures, from literature [26]. Intrathecal hydration was restored by water injection (∼60mL) delivered at the cervical decapitation level to re-inflate and lubricate the intrathecal spaces. An introducer sheath (5-Fr) was inserted into the subarachnoid space, on the posterior surface of the spinal cord, at the level of decapitation on the cervical spinal column (Fig 1, A) and advanced cranially toward the cisterna magna.

**Fig 1:**
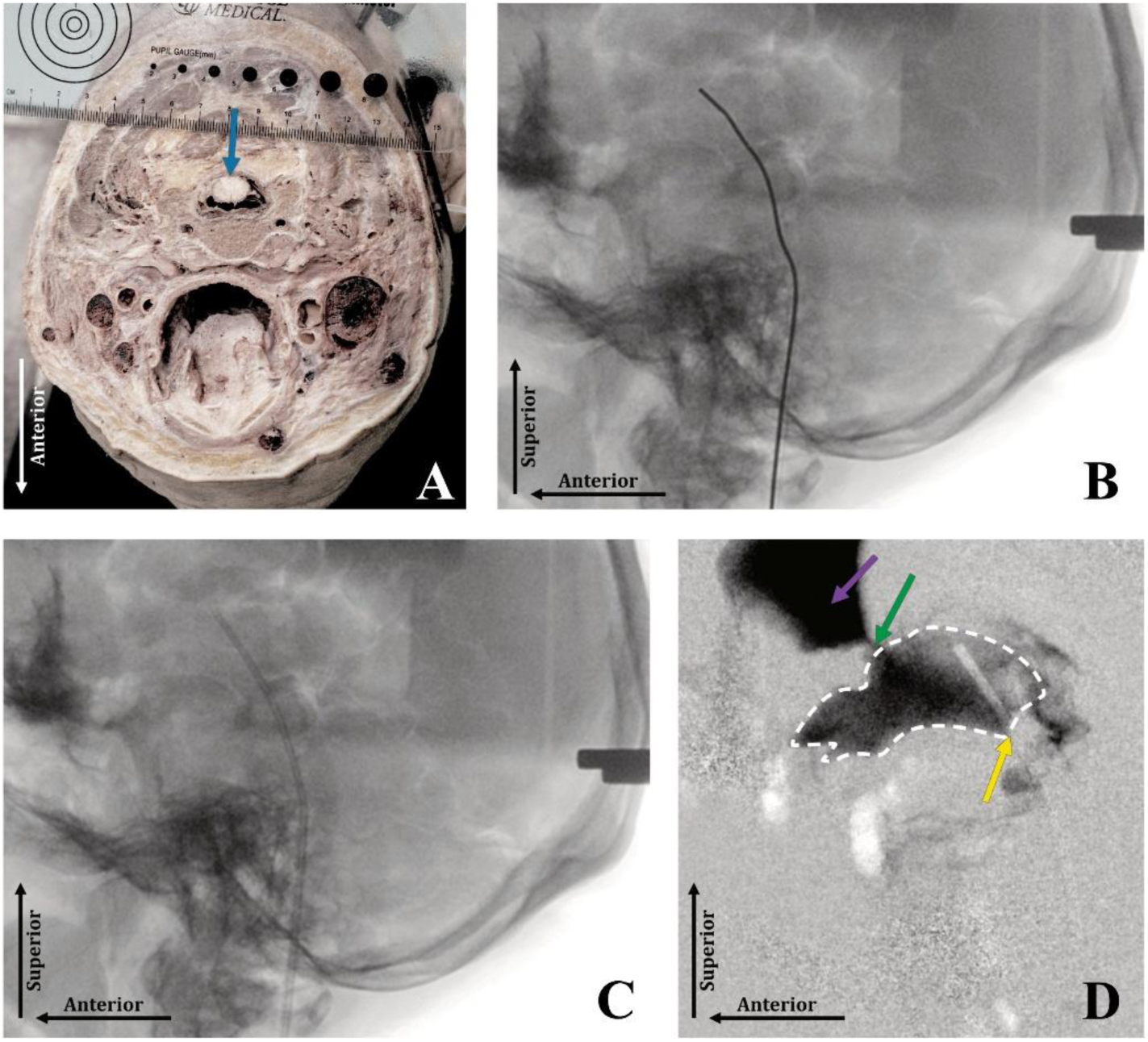
**A.** Presentation of the cadaveric specimen in prone position. The intrathecal access point is shown on the posterior aspect of the spinal cord (blue). **B, C.** Fluoroscopic visualisation of guidewire and catheter (respectively) advancement through the fourth ventricle and cerebral aqueduct into the third ventricle. **D.** Contrast subtraction enhanced third ventriculogram showing third ventricle (white, dotted line), catheter traversing the cerebral aqueduct (yellow), Interventricular foramen (green), and the lateral ventricle (pink).

Under fluoroscopic guidance, a 0.035″ (0.889 mm) guidewire (Nitrex, ev3, USA) was advanced through the introducer, through the subarachnoid space to the level of the fourth ventricle. At this point the 0.035” guidewire was exchanged for a more flexible 0.018″ (0.457 mm), 45° angled-tip wire (Glidewire Gold, Terumo, Japan) in order to navigate the cerebral aqueduct, which progresses at an anterior angular offset of approximately 40°-50° taken from the fourth ventricular midline trajectory. Third ventricular access was defined as catheter tip inside the third ventricular cavity, confirmed using contrast ventriculography. Following successful third ventricular access and catheterisation with a 4-Fr (1.32 mm) catheter (PowerPICC, BD, USA), the 0.018” guide wire was exchanged back to a 0.035” guide wire for improved structural support for repeat catheter exchanges (Fig 1, B). Sequentially larger catheters (4Fr–6Fr PowerPICC, BD, USA) were passed over the in-situ wire to allow a qualitative assessment of resistance (defined in table 1) and maximal diameter accommodation of the cerebral aqueduct (Fig 1, C). Catheter insertion was stopped at 6-Fr (1.98 mm) (as this matched the expected upper boundary of the inner-diameter of the cerebral aqueduct as inspected on MRI imaging), or at the point when a catheter was no longer able to be inserted through the aqueduct pathway, with the last successful catheter insertion recorded for catheter diameter and level of resistance. In the case of specimen 2, an additional 8.5-Fr (2.8 mm) catheter (Attain Command, Medtronic, Ireland) was successfully advanced into the third ventricle. Finally, an Iohexol contrast (Omnipaque 350 mg, GE Healthcare, USA) bolus was administered for visualization via a fluoroscopic contrast subtraction third-ventriculogram (Fig 1, D). To maintain cadaveric specimens for formal dissection, objective post-procedural histopathological assessment was not carried out in this study, and the anatomical dissection protocol was not designed to assess gross anatomical trauma arising from the access procedure.

**Table 1:** Definitions for the qualitative level of resistance noted during procedural catheterisation of the cerebral aqueduct.

| Resistance: | Definition of Resistance: |
| --- | --- |
| None | No resistance felt by operator |
| Minor | Slight resistance felt by operator, no impact on procedure |
| Moderate | Moderate resistance felt on catheter advancement, minimal impact on procedure (slight torquing and/or back-and-forth movement employed to assist advancement) |
| Strong | Strong resistance felt by operator with procedural impacts (required torquing and/or back-and-forth motions to allow advancement) |
| Impassable | Unable to pass catheter through aqueduct |

It should be noted that cadaveric brain tissue specimens are expected to shrink following formalin fixation, with reported cerebral volume shrinkage of 10 – 15% reported in literature [30], [31].

### 2.5. Anatomical Dissection Protocol

Following the third ventricular access evaluation, three cadaver specimens were dissected to visualise and measure the minimally invasive cerebral aqueduct pathway described in 2.4. The goal was to describe and measure dimensions of the pathway and the spatial relationships of the third ventricle to the Thalamus, STN, and GPi. Throughout the dissection procedure, the dimensional measurements were recorded using a scale ruler and digital callipers with these measurements confirmed with post-procedural analysis of digital photographs.

To facilitate access, removal of the calvaria and the occipital bone posterior to the foramen magnum was required. Laminectomy of the upper cervical vertebrae was also performed, with removal of dura and arachnoid mater exposing the posterior aspect of the spinal cord and cisterna magna. Subsequent removal of the cerebellum and roof of the fourth ventricle exposed the rhomboid fossa. Following removal of the brainstem via an axial section through the midbrain, a midline sagittal dissection of the third ventricle was performed. Sagittal sectioning was then extended to separate the left and right cerebral hemispheres entirely. Each hemisphere was then sliced in either the axial or coronal plane. This permitted direct visualisation and measurement of the spatial relationships of key structures (STN, GPi) to the third ventricle.

## 3. Results

### 3.1. MRI assessment

MRI assessment was successfully completed, and the corresponding morphometric data is presented in table 2.

**Table 2:** Average dimensions of third ventricular structures measured from MRI datasets (n = 16).

| Category | Category Description | Rater 1<br>Mean ± SD | Rater 2<br>Mean ± SD | Combined<br>Mean ± SD |
| --- | --- | --- | --- | --- |
| 1 | 3rd Ventricle Minor Axis Length (mm) | 17.90 ± 1.60 | 17.97 ± 1.73 | 17.94 ± 1.63 |
| 2 | 3rd Ventricle Major Axis Length (mm) | 30.82 ± 2.26 | 30.75 ± 1.91 | 30.79 ± 2.04 |
| 3 | 3rd Ventricle Width (mm) | 4.56 ± 0.94 | 4.69 ± 0.91 | 4.63 ± 0.93 |
| 4 | Cerebral Aqueduct Mean Diameter (mm) | 1.60 ± 0.21 | 1.60 ± 0.22 | 1.60 ± 0.21 |
| 5 | Cerebral Aqueduct Length (mm) | 19.38 ± 1.72 | 19.43 ± 1.92 | 19.40 ± 1.81 |
| 6 | Projection Length from Aqueduct Ostia into 3rd Ventricle (mm) | 21.84 ± 1.25 | 22.56 ± 1.52 | 22.20 ± 1.36 |
| 7 | Massa Intermedia Major Axis Length (mm) | 13.45 ± 2.00 | 13.06 ± 1.86 | 13.25 ± 1.92 |
| 8 | Massa Intermedia Minor Axis Length (mm) | 8.86 ± 1.38 | 8.85 ± 1.30 | 8.85 ± 1.31 |
| 9 | Massa Intermedia Contact Area (cm<sup>2</sup>) | 0.84 ± 0.25 | 0.83 ± 0.25 | 0.84 ± 0.25 |
| 10 | Tuber Cinereum Length (mm) | 5.04 ± 0.77 | 5.09 ± 0.74 | 5.06 ± 0.74 |
| 11 | Projection Length from Tuber Cinereum into 3rd Ventricle (mm) | 9.01 ± 1.42 | 9.24 ± 1.20 | 9.13 ± 1.29 |
| 12 | Anterior Length, C1-C2 to 3rd Ventricle (mm) | 99.11 ± 6.34 | 98.57 ± 6.38 | 98.84 ± 6.32 |
| 13 | Posterior Length, C1-C2 to 3rd Ventricle (mm) | 101.66 ± 6.08 | 101.35 ± 5.87 | 101.50 ± 5.95 |
| 14 | Anterior Path, Max. Curvature ($\text{mm}^{-1}$) <sup>†</sup> | $0.07 \pm 0.05$ | $0.07 \pm 0.02$ | $0.07 \pm 0.03$ |
| 15 | Posterior Path, Max. Curvature ($\text{mm}^{-1}$) <sup>†</sup> | $0.20 \pm 0.12$ | $0.18 \pm 0.08$ | $0.19 \pm 0.09$ |
| 16 | Post. Path, Mean Curvature ($\text{mm}^{-1}$) | $0.023 \pm 0.007$ | $0.027 \pm 0.008$ | $0.025 \pm 0.006$ |

\* n = 16 for all categories, <sup>†</sup>Curvature values are the mean ± SD of per-subject maximum curvature along the path centreline.

### 3.2. Feasibility of Access

Trans-aqueduct access to the third ventricle was successful in 5 of 6 specimens (83%). Inability to track the guidewire into the cerebral aqueduct from the fourth ventricle was observed via a perforation event in one specimen, with retrospective analysis suggesting the cause was due to procedural complications compounded by a high likelihood of inconsistent tissue fixation in the region of the quadrigeminal plate. This likely resulted in a reduction in the structural integrity of the trans-aqueduct pathway due to tissue degradation and caused the inability of the guidewire or catheter to track the anterior curvature without penetrating through to the quadrigeminal cistern, especially prevalent during attempts to navigate the anterior curve of the cerebral aqueduct. As such, this event may not be representative of the expected in-vivo tissue behaviour.

Catheter resistance varied across specimens (Table 3). Minor resistance was most common (3/5, 60%) of cases in which access was achieved; moderate resistance was encountered only with catheters ≥1.7 mm in two of five (40%) cases.

**Table 3:**
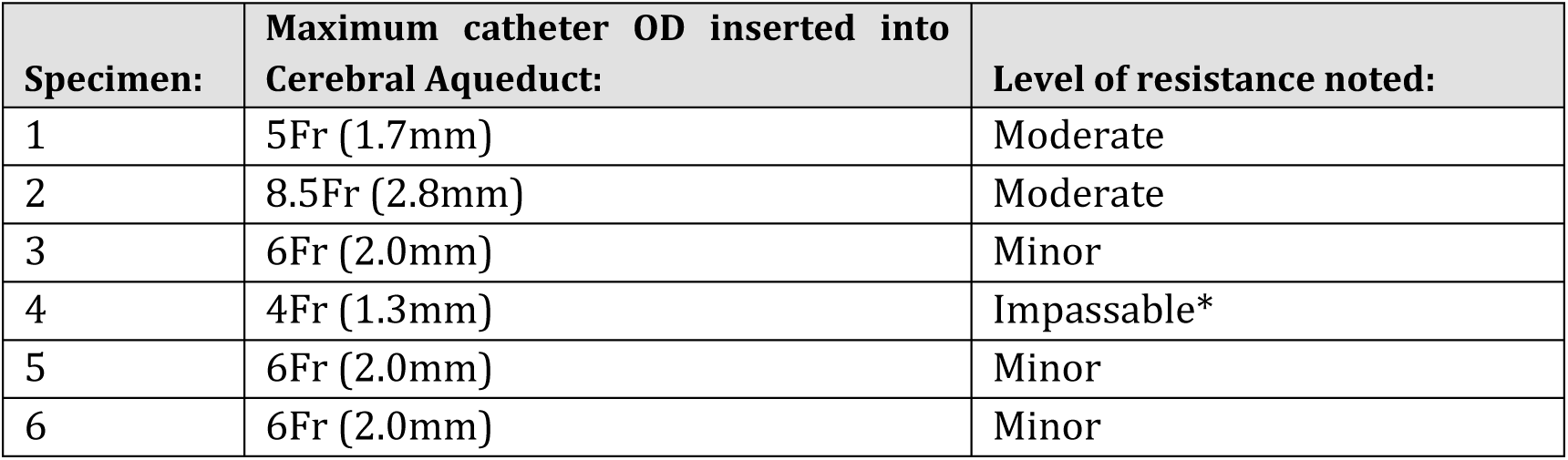
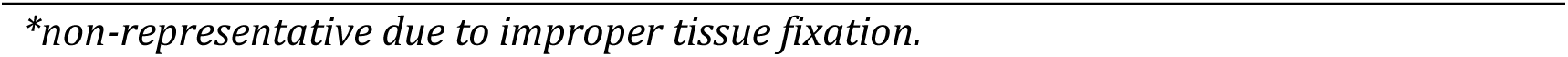
Qualitative assessment of catheter resistance during advancement through the cerebral aqueduct.

| Specimen: | Maximum catheter OD inserted into Cerebral Aqueduct: | Level of resistance noted: |
| --- | --- | --- |
| 1 | 5Fr (1.7mm) | Moderate |
| 2 | 8.5Fr (2.8mm) | Moderate |
| 3 | 6Fr (2.0mm) | Minor |
| 4 | 4Fr (1.3mm) | Impassable* |
| 5 | 6Fr (2.0mm) | Minor |
| 6 | 6Fr (2.0mm) | Minor |

*\*non-representative due to improper tissue fixation.*

The cerebral aqueduct, ventricular cavities, and associated structures were visualised fluoroscopically following contrast injection (Fig 2) for purposes of landmark detection. The procedural time for successful access was approximately 15 to 30-minutes. It should be noted that this procedural duration is for access from the cervical spinal level. A longer procedural duration is anticipated when carrying out lumbar puncture and trans-spinal tracking.

**Fig 2:**
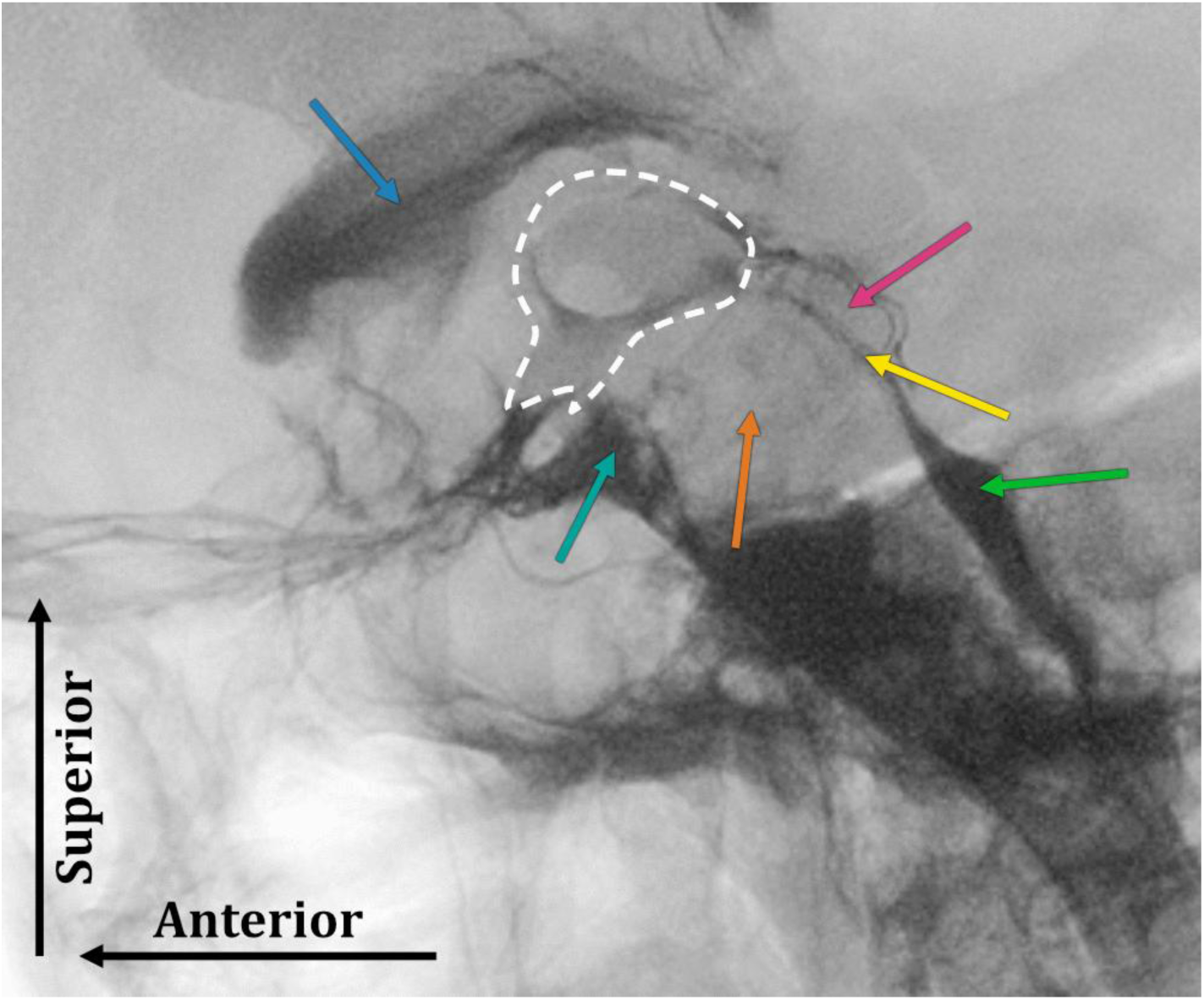
Fluoroscopic image showing contrast delineation of the fourth ventricle (green), cerebral aqueduct (yellow), corpora quadrigemina (pink), third ventricle (white region, approx.), midbrain (orange), interpeduncular cistern (aqua), and lateral ventricle (blue, arrow).

### 3.3. Anatomical Characterisation

The explored anatomical pathway extended from the subarachnoid space surrounding the superior cervical spinal cord, through the cisterna magna, passing into the ventricular system via the foramen of Magendie. From this point, the luminal pathway was followed through the 4^th^ ventricle and cerebral aqueduct to reach the midline third ventricle. Additionally, the anterior brainstem and related cisterns were inspected. Measurements were recorded and features of tortuosity, vascularisation, and anatomical structures that may impede the advancement of interventional access systems were examined as described below.

Within the subarachnoid space the posterior aspect of the spinal cord is covered by the closely adherent pia mater and presented a generally smooth, convex surface over which guidewire and catheter advancement was feasible. Dauleac, C. et al described a median dorsal arachnoid septum extending along the length of the cord in the posterior midline [32]. However, this was not observed during this dissection when the dura and arachnoid were removed from the posterior aspect of the cord. Spinal nerve roots projecting bilaterally from the postero-lateral margins of the spinal cord were separated by a midline longitudinal band approximately 10 mm wide along the posterior surface of the cord. This band provides a space that may allow guidewire and/or catheter systems to advance through the intrathecal space, along the spinal cord midline, with a lowered risk of traumatic interference with spinal nerve roots. The subarachnoid space posterior to the spinal cord in the upper cervical region has been reported as 2.34 mm ± 0.87 mm at the level of the C2-3 intervertebral disc, gradually decreasing to 1.80 mm ± 0.68 mm at C6-7 intervertebral disc level [33]. In the access portion of this study the guidewires used to pass through this space were 0.45 – 0.89 mm in diameter, and catheters up to 2 mm diameter.

At the level of the junction of the spinal cord and medulla oblongata, the subarachnoid space enlarges as the arachnoid bridges over the dorsal aspect of the medulla oblongata to reach the inferior aspect of the cerebellum. This forms the posterior cerebellomedullary cistern (cisterna magna) which contains vascular structures including the vertebral arteries, the posterior inferior cerebellar arteries and the choroid plexus. The lower four cranial nerves – glossopharyngeal, vagus, spinal accessory and hypoglossal – also pass through the lateral aspect of this cistern [34]. In one of three (33%) specimens examined in this study the tonsillomedullary portion of the posterior inferior cerebellar artery (PICA) extended into the cavity volume of the cisterna magna (Fig 3, A). Various morphologies of the cisterna magna have been described with subtypes being significantly influenced by sex. The typical CSF volume in the adult cisterna magna ranges between 0 and 1091.26 mm^3^ ± 277.98 mm^3^, depending on cisterna magna type [35] however, the volume is reportedly smaller in females and the cisterna magna three times more likely to be absent [35].

**Fig 3:**
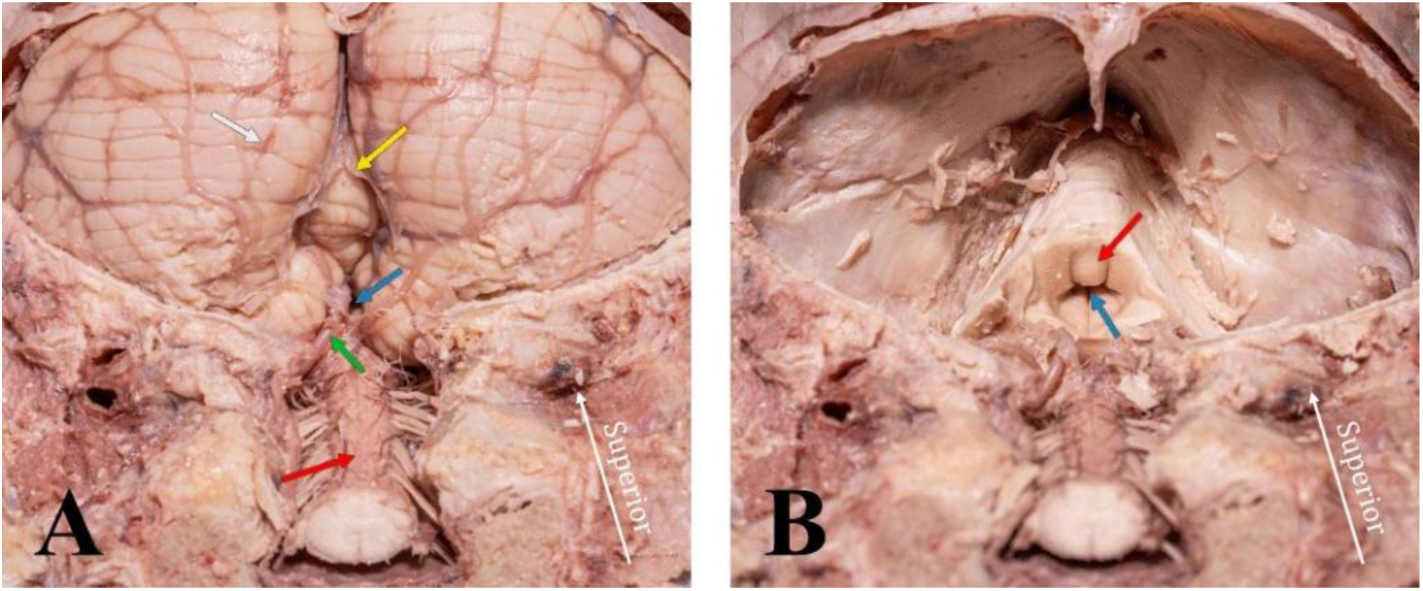
Postero-inferior view of spinal column and skull base dissection, with **3A.** showing the spinal cord (red), median aperture (blue), remnants of the arachnoid mater which enclosed the cistern magna (yellow), the base of the Cerebellum (white), and the tonsillomedullary portion of the PICA (green), and **3B.** with cerebellum removed, showing the interior surface of the fourth ventricular cavity (red), and the rostral opening of the cerebral aqueduct (blue).

The cisterna magna is continuous with the ventricular system via the foramen of Magendie of the fourth ventricle. The ventricular system and foramen of Magendie are lined by ependymal cells with microvilli on their apical surfaces that facilitate the movement of CSF [34]. During the dissection phase of this study, the foramen of Magendie was found to be sufficiently patent across all three evaluated specimens.

Considering the access portion of this study, on entering the 4th ventricle, guidewires were successfully passed through the 4^th^ ventricular cavity over the longitudinal median sulcus thereby minimising risk to underlying cranial nerve nuclei and brainstem pathways. Extending rostrally for the height of the fourth ventricle the cranial-rostral apex of the 4^th^ ventricle and inferior aperture of the cerebral aqueduct was accessed. A predominantly straight trajectory was found extending along the posterior surface of the spinal cord from the cervical level, through the cisterna magna, median aperture (Fig 3, A) and fourth ventricle (Fig 3, B). From a mechanical perspective, the tortuosity of this intrathecal pathway was found to be suitable for guide wire and catheter-based systems.

Exploration of the trans-aqueduct pathway extended through the cerebral aqueduct, the narrow channel which extends through the midbrain, connecting the third and fourth ventricles and facilitating the flow of CSF between the two ventricles. The aqueduct is located between the tectum and tegmentum of the midbrain and is surrounded by periaqueductal grey matter along its length. In adults the dimensions of the aqueduct are typically between 1-2 mm in diameter and 14-15 mm in length [34]. Across all subjects examined in this study, the average diameter of the cerebral aqueduct was observed to be 1.6 mm (SD = 0.20 mm) and the average length (evaluated from MRI) was 19.40 mm (SD = 1.81 mm). The pathway through the aqueduct exhibited a gentle superior-anterior curvature of approximately 45° taken from the midline trajectory of the fourth ventricle and the centreline trajectory of the cerebral aqueduct as it meets the third ventricle, inferior to the posterior commissure.

The third ventricle forms a slit-like cavity of approximately rectangular morphology (Fig 4). The roof is bounded superiorly by the body of the fornix and contains the tela-choroidea with the internal cerebral vein between its layers. The floor is formed by neural structures including the hypothalamus, optic chiasm, tuber cinereum, mammillary bodies, posterior perforated substance and anterior aspect of midbrain tegmentum [34]. The lateral walls are formed by the thalamus and hypothalamus and are joined across the midline by the Massa intermedia (MI) in the majority of people [36]. The oval shaped MI, partially dividing the cavity and connecting the left and right thalami was present in two of three specimens dissected in this study (67%). Table 4 presents morphometric measurement data corresponding to the cadaveric specimens, with Fig 5 indicating the measurement protocol. The anterior – posterior dimension of the third ventricle was measured between the lamina terminalis and the posterior commissure with an average dimension across the three specimens of 27.6 mm (SD = 1.78 mm). The average supero-inferior dimension as measured between the anterior aspect of the mammillary bodies and the supero-most point of the roof of the third ventricle was 19.9 mm (SD = 1.69 mm). The average transverse width of the third ventricle was 5.7 mm (SD = 1.53 mm) when measured from the deepest point within the hypothalamic sulci.

**Fig 4:**
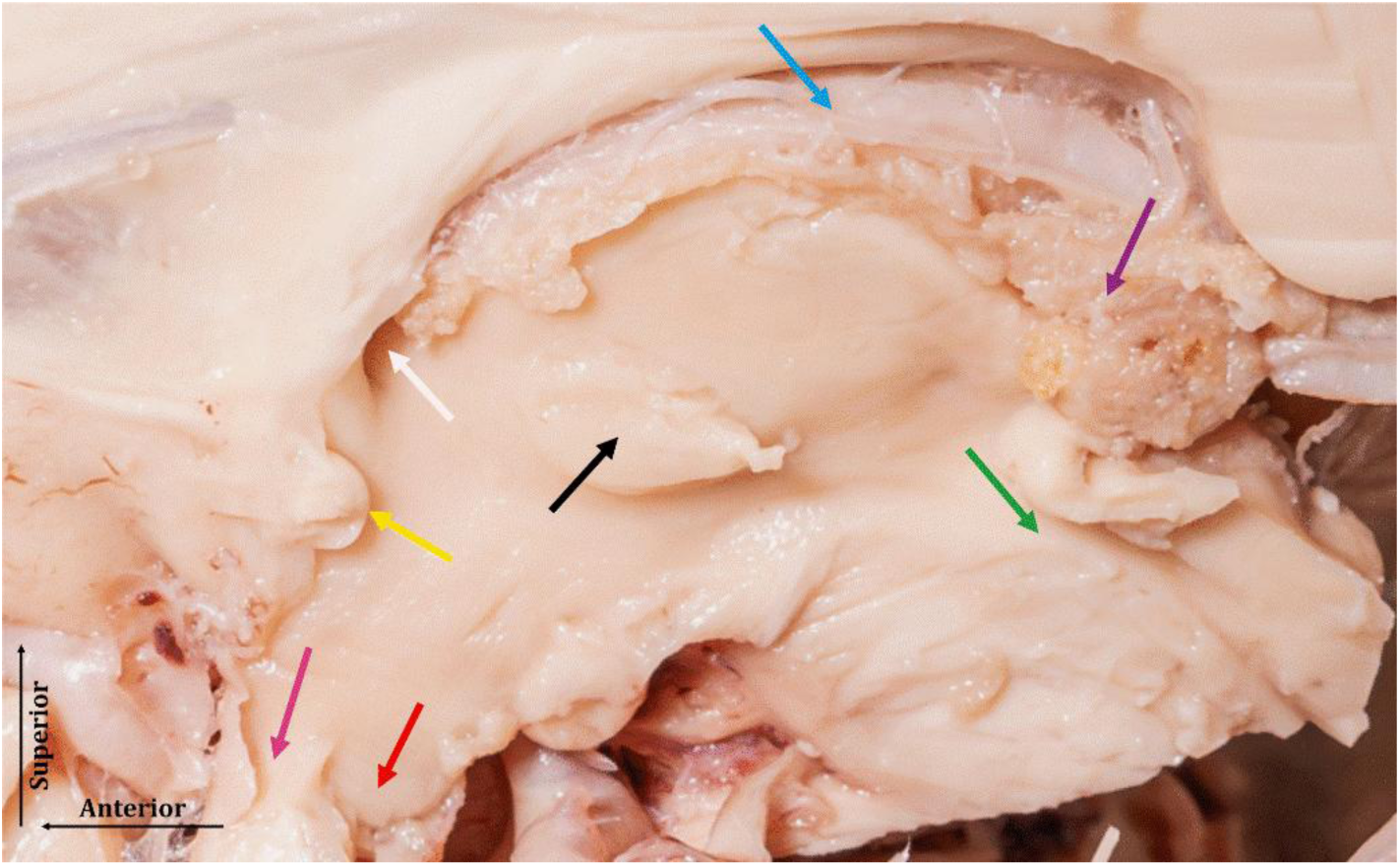
Sagittal midline dissection of the deep brain, viewing the third ventricle showing cerebral aqueduct ostium (green), Massa intermedia (black), infundibular recess (red), supraoptic recess (pink), anterior commissure (yellow), interventricular foramen (white), choroid plexus (purple), internal cerebral veins (blue).

**Fig 5:**
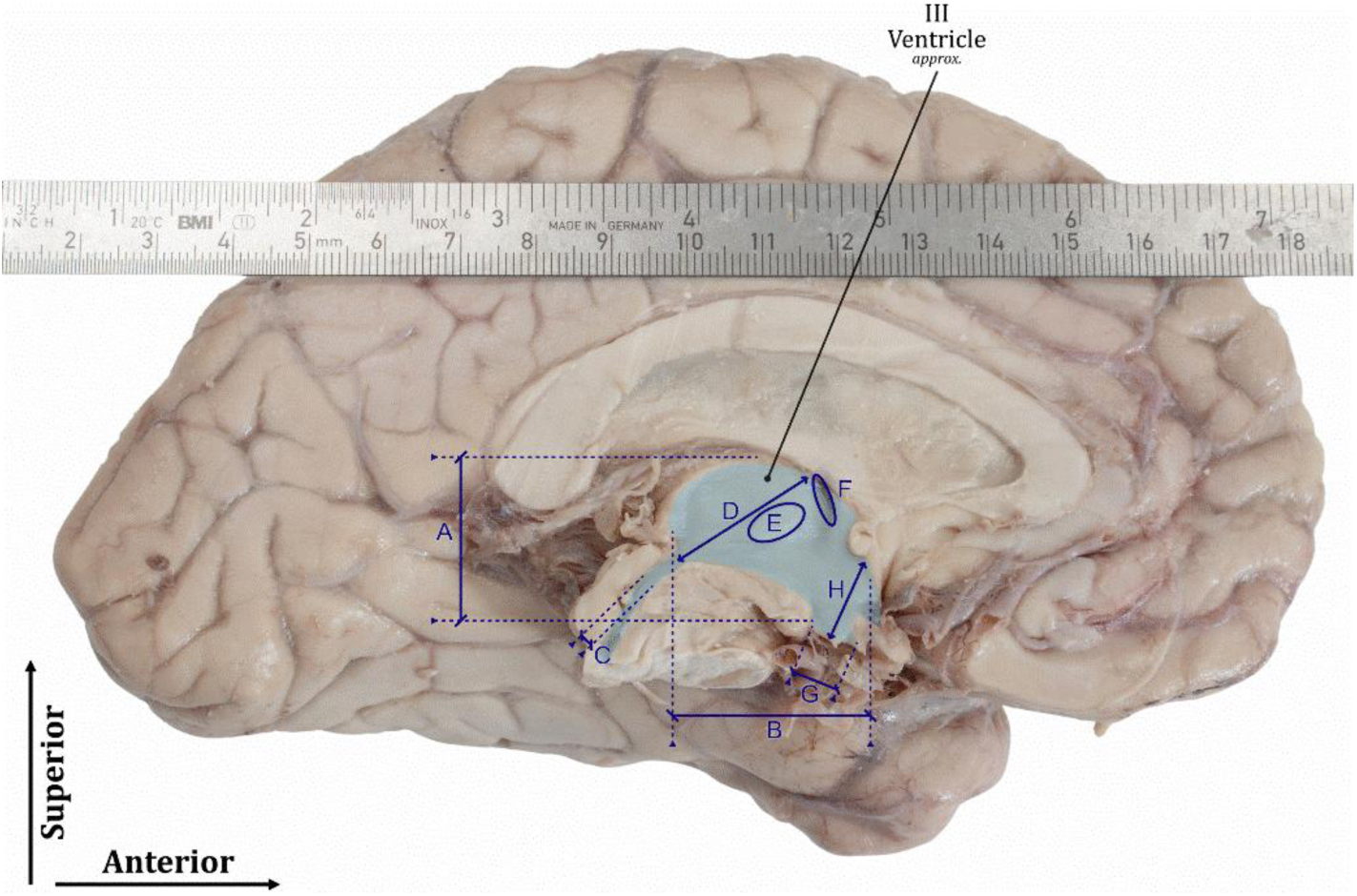
Dissected left cerebral hemisphere showing recorded measurements, A. 3^rd^ Ventricle Minor Axis Length, B. 3^rd^ Ventricle Major Axis Length, C. Cerebral Aqueduct Mean Diameter, D. Projection Length from Cerebral Aqueduct Ostia into the 3^rd^ Ventricle (parallel to sagittal midline), E. Massa Intermedia Minor Axis Length, F. Interventricular foramen - Major Axis, G. 3^rd^ Ventricle tuber cinereum (membranous floor) (co-planar with sagittal midline), H. Projection length from tuber cinereum into the third ventricle (parallel to sagittal midline).

**Table 4:** Dimensional analysis of third ventricular structures in dissected cadaveric specimens (n=3) and MRI subjects (n = 16).

| Measurement | Cadaver - Mean ± SD [mm] | MRI (Combined Observers) - Mean ± SD [mm] | Combined Weighted Mean ± SD [mm] | Coefficient of Variation ($\frac{\sigma}{\mu} \times 100\%$) | Difference, Cadaver - MRI [mm (%)] |
| --- | --- | --- | --- | --- | --- |
| 3<sup>rd</sup> Ventricle Minor Axis Length [A] | 19.9 ± 1.69 | 17.9 ± 1.63 | 18.2 ± 1.75 | 10% | +1.9 (+11%) |
| 3<sup>rd</sup> Ventricle Major Axis Length [B] | 27.6 ± 1.78 | 30.8 ± 2.04 | 30.3 ± 2.28 | 8% | -3.2 (-10%) |
| 3<sup>rd</sup> Ventricle Width | 5.7 ± 1.53 | 4.6 ± 0.93 | 4.8 ± 1.06 | 22% | +1.0 (+23%) |
| Cerebral Aqueduct Mean Diameter [C] | 1.63 ± 0.17 | 1.60 ± 0.21 | 1.60 ± 0.20 | 12% | +0.03 (+2%) |
| Projection Length from Cerebral Aqueduct Ostia into the 3<sup>rd</sup> Ventricle (parallel to sagittal midline) [D] | 21.9 ± 3.02 | 22.2 ± 1.36 | 22.2 ± 1.60 | 7% | -0.3 (-1%) |
| Massa Intermedia Major Axis Length [E] | 8.0 ± 0.11 (n=2) | 13.3 ± 1.92 | 12.7 ± 2.47 | 20% | -5.2 (-40%) |
| Massa Intermedia Minor Axis Length [E] | 4.5 ± 0.49 (n=2) | 8.9 ± 1.31 | 8.4 ± 1.88 | 22% | -4.4 (-49%) |
| Interventricular foramen - Major Axis [F]* | 6.1 ± 0.82 | NA | NA | NA | NA |
| Interventricular foramen - Minor Axis [F]* | 1.9 ± 0.07 | NA | NA | NA | NA |
| 3<sup>rd</sup> Ventricle tuber cinereum (membranous floor) (co-planar with sagittal midline) [G] | 5.7 ± 0.66 | 5.1 ± 0.74 | 5.2 ± 0.75 | 14% | +0.6 (+12%) |
| Projection length from tuber cinereum into the third ventricle (parallel to sagittal midline) [H] | 10.3 ± 0.94 | 9.1 ± 1.29 | 9.3 ± 1.29 | 14% | +1.2 (+13%) |

2 [A-H] see Fig 5, \*Not recorded on MRI dataset

After completing the measurements of the ventricular pathway, the spatial relationships of the third ventricle and the nuclei of the GPi and STN were examined (Fig 6). The basal ganglia are separated from the third ventricle by the nuclei of the thalamus and the hypothalamus. The biconvex shaped STN is located ventral to the thalamus, immediately rostral to the cerebral peduncles of the midbrain and medial to the fibre tracts of the internal capsule. The internal capsule thereby separates the STN from the wedge-shaped internal segment of the globus pallidus (GPi). The GPi is the most medially located nucleus of the nested group which also includes the external segment of the globus pallidus (GPe) and the putamen. These nuclei are separated by the internal and external medullary lamina respectively. The STN and GPi nuclei both form a critical part of the basal ganglia circuitry and therefore play a crucial role in the control and modulation of movement.

**Fig 6:**
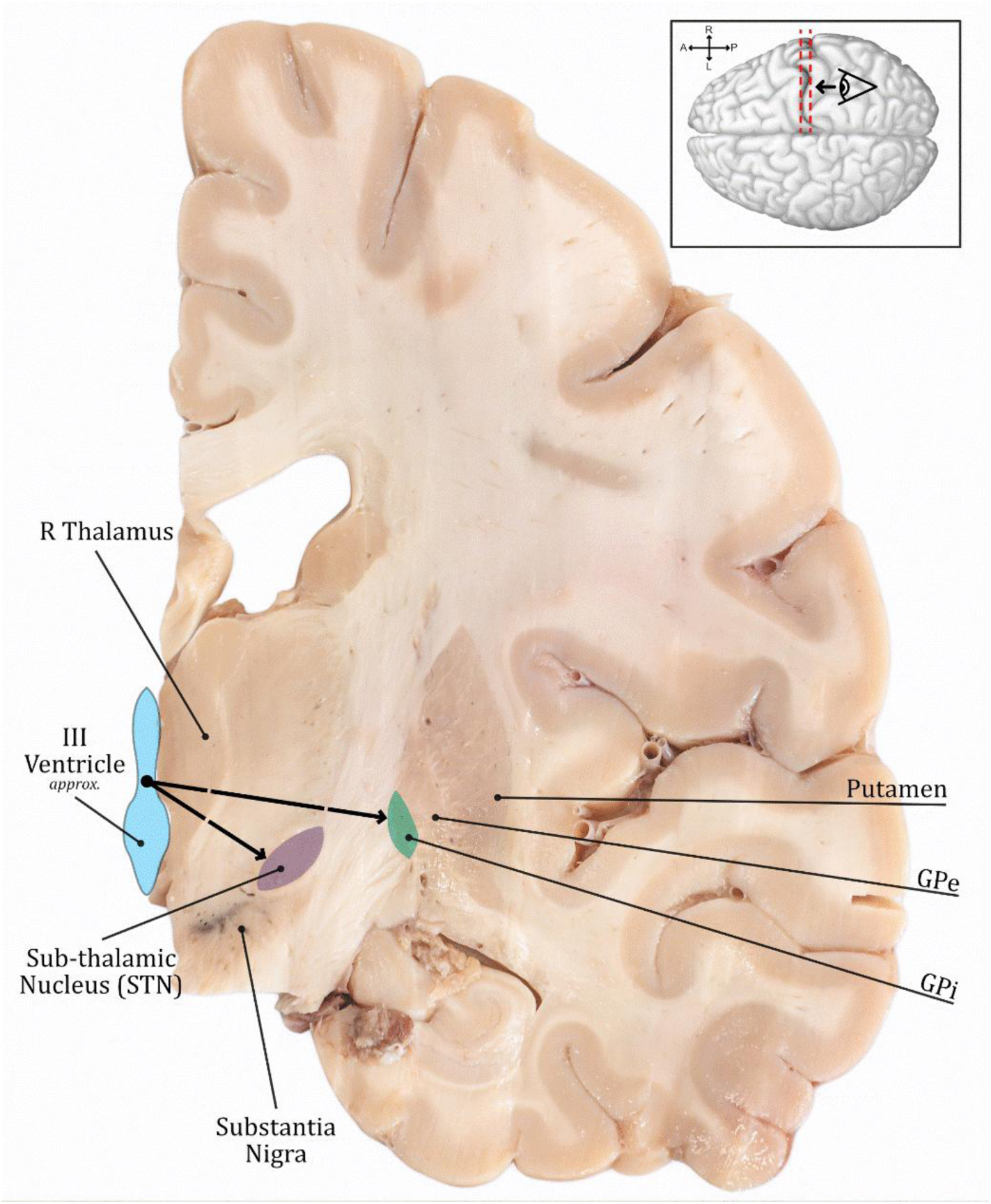
Coronal section of right cerebral hemisphere showing deep brain structures and dimensional relationships of the STN and GPi with the third ventricle centroid.

Layered slicing of the separated cerebral hemispheres in the coronal or axial plane enabled the STN to be identified bilaterally in all three specimens. The distance from the third ventricular centroid to the medial margins of the two nuclei were recorded. The STN was observed to be an almond shaped nucleus which extended obliquely in a superolateral direction along its length. From the midline centroid of the third ventricle, the distance to the medial margin of the left and right STN’s were measured to be approximately 11.4 mm (SD = 1.0 mm). The left and right GPi medial-most borders were observed to occur approximately 20.3 mm (SD = 1.1 mm) bilaterally from the third ventricle centroid.

## 4. Discussion

This cadaveric study demonstrates that trans-cerebral-aqueduct access to the third ventricle is achievable using conventional interventional tools and modern catheter systems. The approach traverses an entirely continuous, and predominantly straight CSF filled pathway (mean curvature κ = 0.025 mm^-1^, SD = 0.006 mm^-1^). The mean maximum curvature κ = 0.19 mm^-1^ (SD = 0.09 mm^-1^), occurring at the junction between the cerebral aqueduct and the fourth ventricle. These curvature values are within the performance limits of typical catheter systems, though careful consideration must be made on the selection of catheter and interventional technologies to ensure their flexural stiffness characteristics and any rigid lengths are suitably managed and the resultant forces applied to surrounding tissue are minimised to help to reduce the risk of penetrative trauma and haemorrhagic-related risks.

The navigation time achieved in this study, requiring approximately 15 to 30 minutes to ascend from the cervical spine to the third ventricle, demonstrates a strictly preliminary observation of the ability to pass the intended pathway from a mechanical standpoint, rather than a clinical representation of procedural duration. In the clinical setting, the procedure would include multiple additional steps including surgical preparation, patient positioning, intrathecal access and navigation over longer spinal pathways, and post deployment verification. Theoretically, if a retrograde, trans-aqueduct deployment approach was translated in-vivo via intrathecal pathway, this method of access to the third ventricle may lead to reductions to the extensive procedural demands associated with conventional DBS implantation, including stereotactic frame alignment, burr-hole craniotomy, trans-cortical trajectory navigation, intraoperative clinical testing, and sub-dermal lead extension tunnelling, managed under a complex asleep-awake-asleep anaesthesia paradigm, which can typically extend up to approximately 6-hours [37].

These potential advantages do, however, remain speculative due to the nature of this study in which only the intra-cranial navigation portion of this intrathecal pathway was observed in fixed, decapitated specimens. Critical translational parameters relating to the real-world full procedural duration, physiological tolerance and burden to the patient, catheter stability under physiological CSF pressure and flow-dynamics, and safety profile relative to conventional DBS require rigorous evaluation through future pre-clinical and in-vivo studies.

Following our analysis of the general dimensionality and geometry of the third ventricle cavity, we can conclude that the volume and arrangement appears suitable to accommodate implantable devices that comprise an overall implanted form factor of up to approximately 5 mm in lateral width and approximately 20 – 30 mm in maximal dimension, which presents more than suitable boundaries for implantable neurotechnology development. Developers must take careful consideration of the variations in the lateral width of the third ventricle and the presence of the MI which may impede available cavity volume to implant technology.

Throughout the analyses, our mean cadaveric and in-vivo MRI data points showed consistent morphological trends across datasets (table 4). It should be noted that the absolute dimensional difference between MRI and direct cadaveric measurements did not vary by more than approximately 3 mm, with the exception of MI measurements, which are discussed below. With respect to the other dimensions which showed limited agreement, being the major and minor axes of the third ventricle, some degree of dimensional difference between cadaver and in-vivo MRI subjects is expected, arising from at least three mechanisms; 1. a significant degree of brain-shift is expected in cadaveric specimens during post-mortem supine storage, 2. formalin fixation is known to cause tissue shrinkage of 10 to 15% [30], [31], and 3. age-related ventricular enlargement is well described in literature with Resnick et. al. describing an annualised ventricular enlargement rate of 1.4 ± 0.1 cm^3^ in elderly subjects [38] and may play a role in the enlarged disparities noted between the cadaveric group (mean age 88.2 years) versus the younger MRI cohort (mean age 48.4 years). These mechanisms are consistent with observations in our data which shows the anterior-posterior axis of the third ventricle to be shorter in the cadaveric versus the MRI cohort (27.6 mm vs 30.8 mm), while the superior-inferior axis dimension was larger in the cadaveric population (19.9 mm vs 17.9 mm), and the lateral width was wider in the cadaveric group (5.7 mm vs 4.6 mm). The present study cannot determine the relative contributions of each of the aforementioned mechanisms in the overall morphometric dataset.

Regarding the differences in measurements of the Massa intermedia (MI) observed between the two datasets; the Siemens Cima.X 3T MRI scans used in the present study were captured with an approximately 0.5 mm voxel resolution. It is theorised that as the left and right thalami protrude medially into the third ventricle volume, pressing together at the level of the MI, the ability to delineate between the thalamic tissue is degraded resulting in over-estimation of the major and minor axes of the MI. This is supported by a moderate-strong correlation (r =-0.6, see S4 Fig. 8) comparing the lateral width of third ventricle cavity and MI area suggesting that narrower third ventricles may have increased contact area of the left and right thalami, making precise MI delineation difficult. This finding is corroborated in prior research evaluating MI anatomy via MRI [36] who noted difficulties in accurately delineating MI borders due to MRI resolution limitations. Again, this finding reinforces the need for development teams to carefully analyse the presence and dimensional arrangement of the MI within the third ventricle to ensure that it is not impeded during access, deployment, or chronic implantation.

Concerning the foramen of Magendie, the observed patency aligns with historical morphometric literature establishing the median aperture as the largest of the three outflow conduits of the fourth ventricle, possessing a mean transverse width of 6.53 ± 1.54mm [39]. This typically large diameter likely enabled successful advancement of our interventional systems from the spinal column into the fourth ventricular cavity, as was observed during the access phase of our protocol. However, the gross morphology of the foramen of Magendie and its immediate structural boundaries are characterised by significant baseline variability [40]. In a clinical setting, transluminal navigation may be heavily complicated by congenital or acquired arachnoid membranes, choroid plexus, asymmetric or descended arrangement of the cerebellar tonsils, and the highly variable PICA branches that frequently line the margins of the aperture. Upon entering the fourth ventricle, the predominantly straight rostral trajectory from the median aperture to the cerebral aqueduct enabled uncomplicated advancement of interventional tools through this cavity, across the dorsal aspect of the longitudinal median sulcus.

The cerebral aqueduct lumen diameter measurements across both the cadaveric and MRI datasets proved consistent (mean = 1.6 mm, SD = 0.20), with excellent MRI inter-rater reliability (ICC 0.94, limits of agreement ±0.15 mm). Paired with the qualitatively assessed ability of the cerebral aqueduct to accommodate catheters up to 2.0 mm in diameter (2.8 mm in a single specimen) without strong resistance, as was observed in the access portion of this study, mechanical feasibility for catheter systems capable of delivering devices to the ventricles via this aqueduct pathway is considered plausible. Tissue-catheter interaction in the present configuration is near-certain in the form of outward radial distension of the aqueduct to accommodate the catheter, as is inferred by the stated aqueduct-catheter diameter mismatch (1.6 mm to 2.0 mm, respectively) which equates to a circumferential strain in the range of 25%. The physiological effects of this distension are not well established and should be a focus of future work exploring larger catheter use. Alternatively, developers should favour catheter sizing that mitigates outward radial stretching on this delicate anatomy. It should be reaffirmed that caution is warranted when interpreting the qualitative nature of the accommodation of catheter systems by the cerebral aqueduct in fixed specimens, as presented in the present study. Absence of significant insertion resistance does not establish an absence of localised trauma. Formalin fixation increases tissue rigidity and alters viscoelastic behaviour, which, when combined with the lack of physiological responses such as bleeding, inflammation, and CSF pressure dynamics within the cadaveric preparation, result in a model that may not be wholly representative of the in-vivo clinical state of tissue, where oversized instruments may cause ependymal tearing, focal shear trauma to the periaqueductal grey matter, or other sub-clinical disruptions. This feasibility assessment should therefore be treated as a basis for future investigations rather than as evidence of clinical safety.

Further, one of the most apparent risks of the cerebral aqueduct as an access pathway to the third ventricle, especially in the context of deploying chronic in-dwelling devices that are intended to remain within the aqueduct, is obstructive hydrocephalus. The choroid plexus within the cerebral ventricles produces CSF at a relatively constant rate of approximately 0.3–0.4 mL/min (∼500 mL/day) [41], and phase-contrast MRI studies in healthy adults have characterised normal trans-cerebral aqueduct CSF mean flow at a proportionate 0.31 ± 0.18 mL/min (net caudal, without respiratory gating), with a cardiac-cycle stroke volume of approximately 41–42 µL per cycle [42], though considerable inter-study variability exists [43]. The Hagen–Poiseuille equation (Eq. 1) describes the pressure differential (ΔP) across a uniform, rigid, cylindrical tube containing an incompressible Newtonian fluid under conditions of steady, laminar flow, providing us with a first-order estimation tool to understand the end-to-end pressure differential required to maintain a steady rate of flow. Critically, ΔP varies inversely with the fourth power of the luminal radius (∝ *r*⁴), indicating a significant sensitivity of the aqueduct to occlusion that must be carefully accounted for in the development and pre-clinical phases of any device intended for this anatomy. Modelling the aqueduct as a cylindrical tube of diameter 1.6 mm and length 19.40 mm (both taken from our averaged MRI dataset, table 2), with a worst case CSF dynamic viscosity *η* = 1.0 × 10⁻³ Pa·s [44] and a mean net flow rate *Q* of 5.17 × 10⁻⁹ m³/s (equivalent to 0.31 mL/min), the baseline pressure differential across the tube is approximately 0.62 Pa (∼0.0047 mmHg). An in-dwelling device spanning the aqueduct and occupying 50% of the luminal cross-sectional area (outer diameter = 1.13 mm in a 1.6 mm aqueduct) converts this lumen to an annular gap. Using eccentric annular Poiseuille flow models, we can compute that for a device which is eccentric to the aqueduct lumen (i.e., outer surface of the member is touching the inner surface of the aqueduct) a pressure differential of approximately 8.9 Pa, or 0.07 mmHg is required to maintain the baseline mean flow-rate, while a perfectly concentric member, forming an annular cross-section with the original lumen, will result in a pressure differential of approximately 21.8 Pa, or 0.16 mmHg to maintain the same flow. When compared to the upper-bound of the normal intracranial pressure (ICP) (∼2000 Pa, 15 mmHg [45]), these increased pressure differentials of 0.07 mmHg and 0.16 mmHg amount to relative increases of just 0.4% and 1.1%, respectively, of the typical upstream intracranial pressure to maintain the normal trans-aqueduct CSF flow rate.

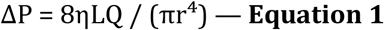

*ΔP = pressure differential (Pa); Q = volumetric flow rate (m³/s); r = luminal radius (m); η = dynamic viscosity (Pa·s); L = tube length (m)*.

It must also be considered that CSF flow through the aqueduct is pulsatile, driven by the cardiac cycle, rather than strictly steady state flow, as is modelled by the Hagen-Poiseuille equations. Employing a Womersley analysis (Eq. 2) allows us to estimate the ratio of transient inertial forces to viscous forces. At a heart rate of 70 bpm, an unobstructed aqueduct lumen of 1.6 mm diameter yields a Womersley number of approximately Wo = 2.17, predicting that the pressure amplitude under pulsatile conditions is approximately 29% higher in the unobstructed lumen versus Hagen-Poiseuille estimates. With an in-dwelling member comprising 50% of the cross-sectional area and creating an annular gap of 0.23 mm, we see a Womersley number of Wo ≈ 0.3 – 0.6, placing flow in this condition within the quasi-steady regime where the viscous effects of the Stokes boundary layer are dominant in the flow dynamics. In this instance, Hagen-Poiseuille annular model estimates represent an accurate approximation of the pressure differential and flow dynamics across the aqueduct where the pressure overhead produced by inertial forces, not captured in the Hagen-Poiseuille model, is relatively small (< 1%).

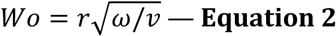

*Wo = Womersley number (dimensionless), r = radius of the lumen (m), ω = angular frequency of the pulsatile pressure (rad/s), ν = kinematic viscosity of the fluid (m2/s)*.

If we model expected peak flow rates achieved during cardiac systole against the above luminal configurations, we can build on our estimates to show that for 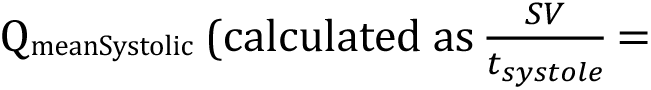 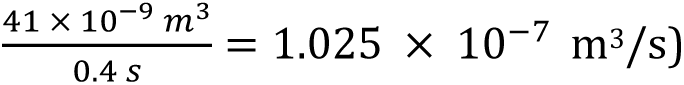 the peak systolic pressure differential required across the aqueduct rises from ∼0.12 mmHg (including the 1.29x Womersley correction) in an unobstructed state, to approximately 1.32 mmHg in the eccentric state and 3.24 mmHg in the concentric state, to maintain the baseline pulsatile flow-rate, resulting from the redistribution of shear stresses along surface boundaries, further resisting flow. Pressure differentials of this magnitude are non-trivial when compared to the baseline pulsatile ICP amplitudes (median ∼2 mmHg [46], abnormal pulse amplitude = sustained ≥ 4 mmHg mean wave amplitude [47]) and may result in reduced caudal pulsatile CSF flow, tri-ventricular CSF accumulation, and increased ICP – each consistent with the mechanism for obstructive hydrocephalus [48]. We can understand from the above illustrative modelling that engineers must continue to strenuously consider the delicate flow dynamics through any fluid path intended for indwelling technologies, such as the cerebral aqueduct, to ensure that the risks of impacting physiological processes are mitigated.

Additionally, for any indwelling device chronically situated within the cerebral aqueduct, engineers and development teams must carefully consider the mechanical interactions between the implant and the surrounding neural structures — specifically the ependymal cell lining and adjacent parenchyma. Chronic CNS implants are known to elicit a foreign body response characterised by microglial activation and astrogliosis [49], which not only impairs local neuronal function but may alter the mechanical properties of the tissue-device interface over time, potentially increasing adhesion and complicating device explant. Repeated micromotion of an implant against the ependymal lining is likely to exacerbate this glial scarring response. Furthermore, outward radial forces, such as those generated by a stent or an oversized shunt, risk localised ischemia and neurological deficit if they impinge upon functionally eloquent structures. This concern is particularly acute with respect to the periaqueductal grey matter (PAG), a structure critical to pain modulation, descending motor control, and autonomic regulation, where even focal ischaemic injury may produce significant and potentially irreversible neurological sequelae. Notably, endoscopic aqueductoplasty, and aqueduct stenting has been explored clinically for the management of cerebral aqueduct stenosis and trapped fourth ventricle [26], with transient ophthalmoparesis (resolving within 2-months) the most common complication reported during aqueductoplasty with, or without stent placement.

In any case, the foramen of Magendie - fourth ventricle - cerebral aqueduct pathway is a compelling pathway through which to gain access to the third ventricle, however, extreme caution must be used when considering the impact of any such technology intended for access through this pathway and its potential interference with surrounding parenchyma and the critical physiological function of the aqueduct in maintaining caudal CSF flow.

Potential clinical morbidity and mortality risks for trans-aqueduct, third ventricular access are numerous and must be weighed against the expected benefit of undertaking such a procedure. Some potential risks include, but are not limited to, trauma to neural structures and or delicate lining of the ventricle and aqueduct cavities arising from inability to track the patent midline of the intrathecal and aqueduct pathways; interference with brainstem and/or cerebral structures including direct trauma, outward radial pressure injuries occurring from systems or devices that are oversized for the anatomy; occlusion of any component the CSF pathways resulting in hydrocephalus; and, trauma to the delicate choroid plexus and/or vascular anatomy, which extend into the luminal cavities and may interact with endoluminal systems as they navigate the pathway. These risks are closely linked to the guidewire and catheter systems being implemented and may be mitigated through thoughtful development of endoluminal systems that are configured to reduce penetration and trauma risk, and which are optimised for stable midline tracking through the delicate anatomy. Anatomical variability in aqueduct diameter and ventricle morphology will likely necessitate thorough pre-operative planning (i.e., MRI/CT) to define the cerebral aqueduct pathway, paired with real-time, intraprocedural image guidance (i.e., fluoroscopy) to ensure interventional systems do not deviate from the defined pathway.

From a neurotechnology perspective, the third ventricle lies adjacent to bilateral deep nuclei that are proven targets in established neuromodulation therapies. In the specimens evaluated in this study, the medial margins of the STN and GPi were located approximately 11.4 mm to 20.3 mm, respectively, from the centre of the third ventricle. These dimensions agree with published literature of larger population datasets, which show measurements 7.9 – 12.1 mm to the medial STN border [50] and 16.0 – 25.45 mm to the posterior ventrolateral GPi [51], taken from the mid-commissural point in both instances. The primary relevance of this proximity is as a design parameter that defines the stand-off distance through which an intraventricular system would be required to act. This measurement is further affected by the intended positioning of a device within the ventricle, which would likely be against the lateral wall of the cavity, resulting in a reduction in this stand-off distance, equal to half of the third ventricular lateral width (2.8 mm in the cadaver cohort of the present study). These measurements are derived from a small, fixed, aged cadaveric sample population and should be confirmed through further in-vivo studies. The ability of any neuromodulation systems or technologies to achieve selective, therapeutic effect across the stand-off distances remains out-of-scope in the present study.

Future work should focus on in vivo validation in animal models, including dynamic imaging to assess CSF flow effects and neurological safety, development of atraumatic catheter systems optimised for trans-aqueduct navigation, and full investigation as to the procedural benefits provided by the present methods. Further, it goes without saying that engineering teams must place critical importance on the strenuous development and validation of device functionality for long-term implantation including deployment methods, stabilisation within the anatomy, and therapeutic benefit of neurotechnologies intended for chronic implantation within the ventricular cavities.

This study has limitations and drawbacks; 1. The age range of cadaver specimens is constrained by the availability of body donors resulting in an observed inter-cohort age disparity in the present study (88.2 years, cadaveric group; 48.4 years, MRI group). Physiological brain atrophy and compensatory ventricular enlargement amongst older subjects may alter luminal compliance and dimensionality relative to younger populations. These age-related effects bear on the interpretation of feasibility in the present study, and the small cadaveric cohort limits confidence in comparative assessments. Developers should reference in-vivo imaging of intended patient populations when designing medical systems, rather than rely on cadaveric datasets. 2. The differences between fixed cadaveric tissue and living tissue are well understood with respect to lack of fluid pressures, tissue degradation, tissue stiffness, and fluid flow, which all impact the ability to gain reliable mechanical measurements from donor specimens. 3. The postmortem shift of brain tissue, including following gross dissection, in cadaveric specimens introduces unreliability into dimensional measurements. 4. The resolution limitations of MRI systems introduce a level of error make accurate delineation of small neural structures difficult, necessitating the employment of multiple observers to improve the reliability of measurements –as was carried out in this study. 5. The scale used to qualitatively determine catheter resistance within the anatomy is subjective to the operator and may limit the translational fidelity of the reported outcomes. 6. All surgical procedures were performed by trained biomedical engineers rather than clinically qualified physicians, which may affect the translational applicability of the surgical outcomes reported. 7. The anatomical dissection portion of the present study was performed on a small specimen subset (n=3), and findings from this portion of the study should be interpreted a descriptive anatomical observation rather than representative of broader population. 8. The clinical status of the MRI cohort was anonymised prior to supply of the datasets. Although no gross anatomical abnormalities were observed, occult pathology, including sub-clinical atrophy and mild enlargement of the ventricles and cerebral aqueduct, have not been evaluated and may have influenced the morphometric values presented in this study. Future studies should prospectively recruit candidate datasets with confirmed neurological health status.

## 5. Conclusion

The present findings show that in the cadaveric setting, trans-aqueduct access is technically feasible from a mechanical standpoint and may enable a minimally invasive route to the third ventricle cavity. Prior to applying any of these findings to clinical practice, extensive and explicit preclinical and clinical evaluation is required to further determine procedural safety, anatomical and physiological compliance, and therapeutic efficacy of any technologies intended for this pathway.

## Data Availability

All data produced in the present study are available upon reasonable request to the authors

## Acknowledgements

The authors sincerely thank those who donated their bodies to science so that anatomical research could be performed. Results from such research can potentially increase humankind’s overall knowledge that can then improve patient care. We also acknowledge the University of Melbourne Department of Anatomy, notably James Heywood for laboratory support and guidance.

## Supplementary

*S1 Fig. 7: Age demographic data of Cadaveric (n=6) versus MRI (n=16) population dataset.*

*S2 Table 6: Inter-rater correlation and agreement data.*

*S3 Table 7: Raw data for MRI and Cadaver subjects.*

*S4 Fig. 8: Correlation plot showing comparison of third ventricular width versus Massa intermedia area, as measured on MRI.*

*S5: Bland-Altman plots and reference data for MRI analysis.*

