## Supplementary material for "Mechanical feasibility of retrograde trans-aqueduct access to the third ventricle: a human cadaveric and MRI morphometric study": HREC Study Exemption Confirmation Letter (from another version of this preprint: version 1, 2026-04-21)

### Office of Research Ethics and Integrity

THE UNIVERSITY OF MELBOURNE

#### Exemption Acknowledged

**Reference Number:** 2026-36108-79208-1
**Project ID:** 36108
**Project Title:** Morphometric analysis of anonymised human cranial MRI imaging

Dear MR Michael Haines,

This email is to acknowledge receipt of your application for Human Ethics Approval Exemption **36108 Morphometric analysis of anonymised human cranial MRI imaging**, which has been granted in accordance with Sections 5.1.16 - 5.1.17 of the National Statement on Ethical Conduct in Human Research.

If the research is changed such that any of these criteria no longer apply, this exemption will cease to be valid and ethics approval must be sought.

If you have any queries concerning this exemption, please contact us via ServiceNow.

Kind regards,

Human Research Safeguards
Office of Research Ethics and Integrity | Research, Innovation & Commercialisation
The University of Melbourne, Victoria 3010, Australia

**Submit an enquiry to the Human Research Safeguards team**

*This email and any attachments may contain personal information or information that is otherwise confidential or the subject of copyright. Any use, disclosure or copying of any part of it is prohibited. The University does not warrant that this email or any attachments are free from viruses or defects.*

*Please check any attachments for viruses and defects before opening them. If this email is received in error, please delete it and notify us by return email.*
